# From Research to Impact: Assessing a Decade of CDC’s Public Health Science by Topic Area, 2014-2023

**DOI:** 10.1101/2025.03.07.25323572

**Authors:** Joy Ortega, Martha Knuth, Victoria E. Dunkley, Christie Kim, Elissa Meites, Brian B. Yoo, Katlyn Wainwright, Bao-Ping Zhu, Mary G. Reynolds

## Abstract

**Objectives:** This study provides an objective, in-depth overview of a large body of science output addressing public health. We apply topic modeling and bibliometric tools to explore the relevance and impact of a decade of CDC-authored publications.

**Methods:** We identified 34,104 scientific publications from 2014-2023 with ≥1 CDC-affiliated author using Science Clips, a CDC library database. We applied a large language modeling framework using BERTopic to publication titles and abstracts to identify public health topic themes. We obtained data from Altmetric, Dimensions, and BMJ Impact Analytics for these publications to bibliometric indicators. We assessed the percent with attention, academic citations, and policy citations using appropriate publication year ranges. We assessed the median Altmetric attention score, median academic citations, and the percent with policy citations for publications by topic area.

**Results:** Of publications from 2014-2020, 95% were cited by academic papers and 52% were cited in clinical guidance or policy. Of publications from 2014-2023, 84% garnered online attention. CDC-authored publications clustered into 46 public health topic themes. Among these, fungal infections had the highest median number of academic citations (36.5), mining safety and health had the highest proportion of papers with policy citations (92.5%), and substance abuse or opioids received the highest median public attention (Altmetric Attention Score = 14). Nearly a third of topics ranked highly (in the top 5) for at least one bibliometric indicator.

**Conclusions:** Publications in this collection addressed an array of public health topic themes and demonstrated resonance within academic and policy arenas as well as with the public.

**SUMMARY:** *What is the current understanding of this subject?:* Public health science should address strategic priorities and translate to public health impact. CDC’s scientists publish over 3,400 articles per year on average, making it difficult to summarize the breadth of topics covered by these articles and how they affect downstream health outcomes.

*What does this report add to the literature?:* We used cutting-edge large language modeling techniques to categorize CDC-authored publications into 46 topic themes. These topic themes span both infectious and non-infectious disease, and cover topics involved in strategic priorities like emergency response. We assess simple indicators like academic citations, policy citations, and media attention by topic area to show that all topic themes have had measurable impact. Across all topic areas, CDC-authored publications are also highly cited in policy and clinical guidance, an indicator of translation to public health impact.

*What are the implications for public health practice?:* Public health research programs can use advances in large language modeling as well as simple indicators of reach and impact to better understand whether their publications address priority topics and translate to improving health outcomes. This overview of CDC research additionally promotes transparency about the activities of the nation’s foremost public health agency.

## INTRODUCTION

High-quality scientific evidence can inform public health policy and practice.^1^ The Centers for Disease Control and Prevention (CDC) within the U.S. Department of Health and Human Services is a science-based, data-driven, government agency with a mission to protect public health.^2^ Manuscripts with CDC-affiliated authors are subject to rigorous internal review to ensure high quality before submission to journals.^4,5^

CDC scientific publications are expected to address strategic priority areas, including emergency response.^6^ Therefore, there is a programmatic need to evaluate whether scientific publications are addressing priority topics and reaching appropriate audiences. A variety of bibliometric methods exist to evaluate reach and impact, each of which has strengths and weaknesses.^7^ Categorizing publications by topic can be labor- and time-intensive, involving manual review and updates to search terms as scientific fields evolve. These challenges are amplified when evaluating a large body of publications from a large organization such as CDC.

To explore the scientific impact and topic areas of CDC’s scientific publications from 2014-2023, we developed a concise set of bibliometric analyses as indicators of possible impact in conjunction with application of a large language modeling technique. Our intent was to identify opportunities to increase scientific impact and promote strategic science at the agency.

## METHODS

We conducted our bibliometric analysis using the BIBLIO checklist.^8^

### Data Sources

We identified scientific papers published from January 1, 2014–December 31, 2023 with ≥1 CDC-affiliated author using CDC Science Clips, a public-facing database maintained by the Steven B. Thacker CDC Library (accessed May 3, 2024).^9^ For publications with a digital object identifier (DOI) or a PubMed identifier (PMID), we obtained Altmetric Attention Scores, the number of Dimensions citations, and the number of policy citations from Altmetric (explorer.altmetric.com; Digital Science, London, UK; accessed July 2, 2024). Additionally, for these same publications, we obtained the number of policy and clinical guidance citations from BMJ Impact Analytics (BMJ and Overton, London, UK; accessed July 2, 2024). Using Microsoft Excel 365 (Microsoft, Seattle, WA), we deduplicated these records based on Altmetric URL (for Altmetric data) and DOI (for BMJ Impact Analytics data). We matched these impact data back to our Science Clips records, further deduplicating Altmetric records that referenced the same publication.

### Topic Modeling

To identify publication topic areas, we used the BERTopic library (version 0.16.0).^12^ BERTopic finds topic clusters using pre-trained large language models (LLM) to create embeddings. We embedded publication titles and abstracts using the SPECTER LLM, which was trained using titles, abstracts, and co-citation networks of scientific publications.^13^ Using this “off-the-shelf” embedding model meant we did not have to fine-tune or train a model from scratch to cluster publications.

For dimensionality reduction, we used the Uniform Manifold Approximation and Projection (UMAP) method.^14^ To cluster the results of our dimensionality reduction, we used Hierarchical Density-Based Spatial Clustering of Applications with Noise (HDBSCAN).^15^ For these steps, we determined parameters that optimized three equally weighted measures: coverage, or the percentage of publications assigned to clusters; the relative validity score, which assesses cluster density as a measure of quality;^16^ and evenness, or the normalized difference between the number of publications in the largest and smallest clusters (eText in the Supplement). We identified optimal parameters using a grid search of 3,600 unique combinations of UMAP parameters (number of neighbors, number of dimensions, and minimum distance) and HDBSCAN parameters (minimum cluster size and minimum samples), while holding the initial random state of UMAP constant. For dimensionality reduction with UMAP, we used these parameters: number of neighbors=25, number of dimensions=50, minimum distance=0.^14^ For clustering with HDBSCAN, we used these parameters: minimum cluster size=175, minimum samples=10. The final model, which was the highest scoring model for this grid search (overall score=0.72), gave us a coverage of 78% (n=26,548), relative validity score of 0.50, and an evenness of 0.89. By the end of these steps, publications were either assigned to a topic cluster or unassigned to any cluster.

The remaining steps of the pipeline identify keywords associated with each cluster. For the vectorizer step, which counts the frequency of “tokens” for all words present in each cluster, we used CountVectorizer with English stop words, a minimum frequency of 2, and an n-gram range of (1,3). We used the default settings for the c-TF-IDF step, which identifies the tokens that appear more frequently in each cluster as the cluster keywords. Finally, we added a KeyBERT-inspired step at the end of the pipeline to produce better keywords; this compares the keywords from the c-TF-IDF step to representative publications from the cluster to fine-tune the chosen keywords further. By the end of these steps, each topic cluster was described using a list of 10 representative keywords and 3 representative publications. To visualize our clusters, we also used a UMAP model with number of dimensions=2 but retained the clustering results and other UMAP parameters from the full-size model.

### Model Quality Assessment

We employed two stages of review to assess the quality of the topic clusters discovered by our topic model (eTable 1 in the Supplement). Our review process included 8 reviewers from CDC staff: 4 senior scientists, 1 subject matter expert (SME) in healthcare quality, 1 SME in global health, 1 librarian, and 2 fellows. In the first stage, four reviewers created short topic labels that described each cluster. Two reviewers assigned topic labels based on the representative keywords, while two reviewers assigned topic labels based on the publications contained within the cluster. These four reviewers then met to reconcile and create provisional topic labels for each cluster. In the second stage, four additional reviewers were involved. We randomly divided the clusters in half and chose 10 random publications from each cluster. For each of the two sets of randomly selected papers (n = 230; 10 from each of 23 clusters), two reviewers were tasked to blindly assign the topic labels created in the first stage to each paper. For the first set of clusters, mean concurrence between the model and human reviewers was 78% (reviewer A: 77%, n=177; reviewer B: 80%, n=184), while the concurrence of the human reviewers with each other was 80% (n=187). For the second set of clusters, mean concurrence between the model and human reviewers was 75% (reviewer C: 73%, n=169; reviewer D: 77%, n=178), while the concurrence of the human reviewers with each other was 84% (n=193). Although significance testing was not conducted, the human reviewers’ concurrence with each other was comparable to their concurrence with the topic model. We also used this stage to inform revisions to topic labels to better reflect the publications in each cluster (eTable 1 in the Supplement).

### Science Impact Metrics

We analyzed metrics for three areas of scientific impact – academic citations, policy citations, and attention – based on data from Altmetric and BMJ Impact Analytics.

Academic citations are a traditional measure of science impact that demonstrate influence and reach of an article’s scientific argument or evidence within an academic field. To assess academic citations, we used the number of Dimensions citations for each publication, available through Altmetric Explorer. Dimensions is another bibliometric tool that tracks academic citations (dimensions.ai; Digital Science, London, UK). The metrics we report based on academic citations include the percent of CDC-authored publications between 2014–2020 that had received any academic citations and the median number of academic citations in each specific public health topic theme during the same period.

Policy citations can measure the uptake and use of an article’s scientific evidence to affect new policy changes or guidance. To assess this measure, we used the number of Altmetric policy mentions and the number of policy or clinical guidance citations in BMJ Impact Analytics for each publication. The metric we report based on policy citations is the percent of all CDC-authored publications between 2014–2020 that had received any policy mentions in Altmetric or policy citations in BMJ Impact Analytics. We also report this metric by public health topic theme.

Citations can take time to accumulate after publication of scientific work. Therefore, when we conducted our analyses in 2024, we chose to limit our data set to publications between 2014-2020 for academic and policy citation metrics. This decision was justified by evaluation of the mean and distribution of the number of academic and policy citations by publication year from our data sources, which appeared stable for 2020 publications (eFigure 1 in the Supplement). This finding is consistent with previous literature showing that both academic and policy citation counts tend to stabilize around 2-3 years post-publication.^18–20^

**Figure 1.**
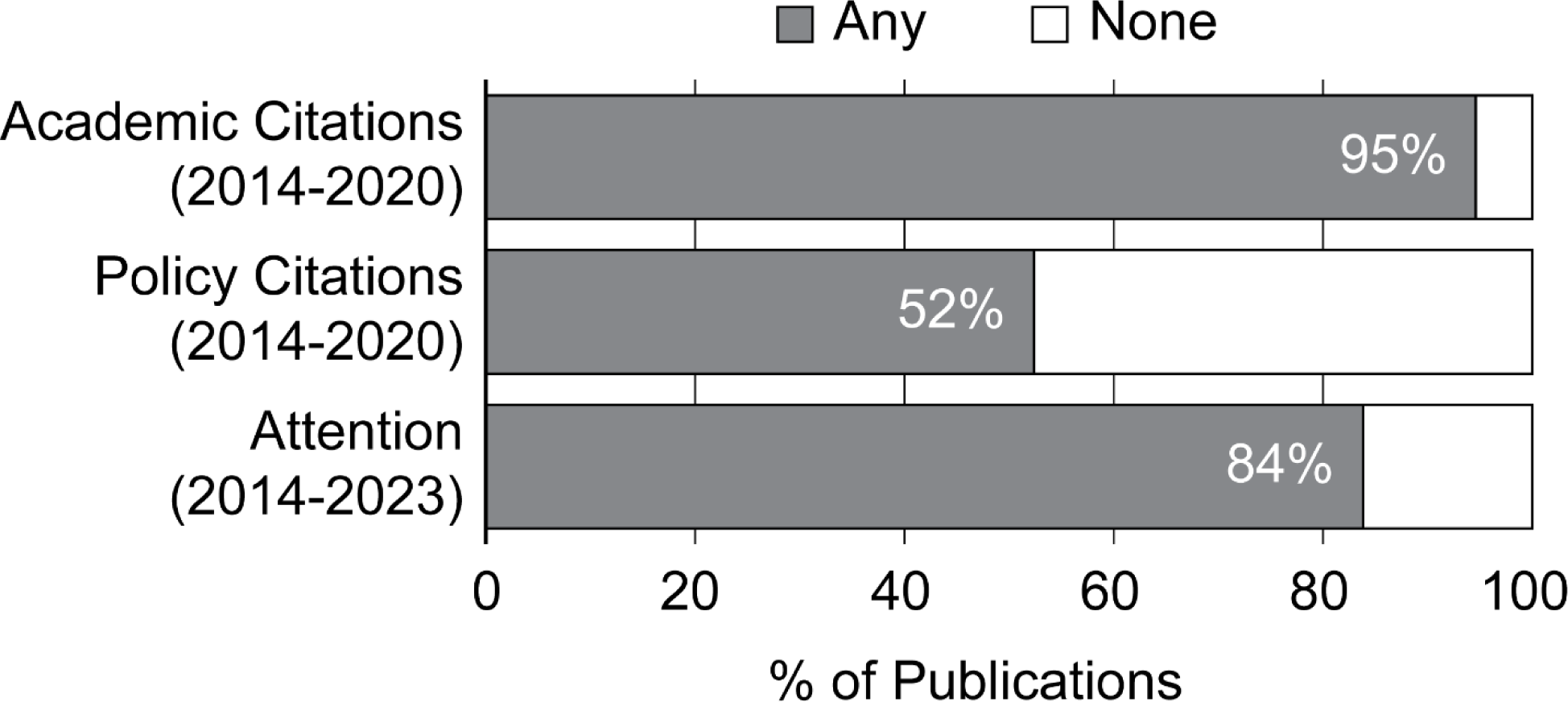
Percent of CDC-authored publications receiving any academic citations, policy citations, and attention during the analytic period. Analyses were conducted in 2024. Academic and policy citation rates typically take approximately 3 years to accumulate (see eFigure 1 in the Supplement), so for these measures, only CDC-authored publications from 2014-2020 (n = 24,208) were considered. CDC-authored publications (2014-2023) were considered to have received attention if they received an Altmetric Attention Score > 0 (n = 34,104). Alt Text: Bar chart reporting the percent of publications receiving any academic citations, policy citation, or attention.

Attention indicators can assess whether information about an article is disseminated through outlets including news media, social media, online forums, and others. To assess attention as a potential indicator of impact, we used the Altmetric Attention Score for each publication, available through Altmetric Explorer. The metrics we report based on attention include the percent of all CDC-authored publications that received attention and the median Altmetric Attention Score for CDC-authored publications in each specific public health topic theme. We defined publications receiving attention as those with an Altmetric Attention Score > 0.

This activity was reviewed by CDC, deemed not research, and conducted consistent with applicable federal law and CDC policy.

## RESULTS

From 2014-2023, CDC authors published 34,104 scientific journal articles indexed in Science Clips, or 3,410 per year on average (range: 2,963 – 3,591). We obtained data from Altmetric for 33,987, and from BMJ Impact Analytics for 14,121, of these journal articles.

CDC-authored publications in this decade had measurable academic, policy, and attention impact (Figure 1). Of the 24,208 publications from 2014–2020 with sufficient time to accrue academic and policy citations, 94% (n=22,896) received at least one academic citation and 52% (n=12,681) received at least one policy citation by the time of our analysis. Of all 34,104 publications from 2014–2023, 84% (n=28,595) received attention indexed in Altmetric.

CDC-authored publications were categorized by our topic model into 46 distinct public health topic themes (Table 1). Topic themes clustered broadly into areas representing infectious diseases, sexually transmitted infections, and chronic health conditions (Figure 2). Topics towards the center of the topic map (suggesting similarity with other topics) included those related to public health practice and capacity, global health security and preparedness, and global emergency response. The largest three topic themes (indicating largest numbers of publications) were related to respiratory illnesses or vaccination (n=3,828, 11.2% of total publications), sexually transmitted infections (n=2,931, 8.6%), and occupational safety and health (n=903, 2.6%). Our model also detected small topic themes including environmental health (n=175) and pandemic influenza (n=177).

**Table 1.**
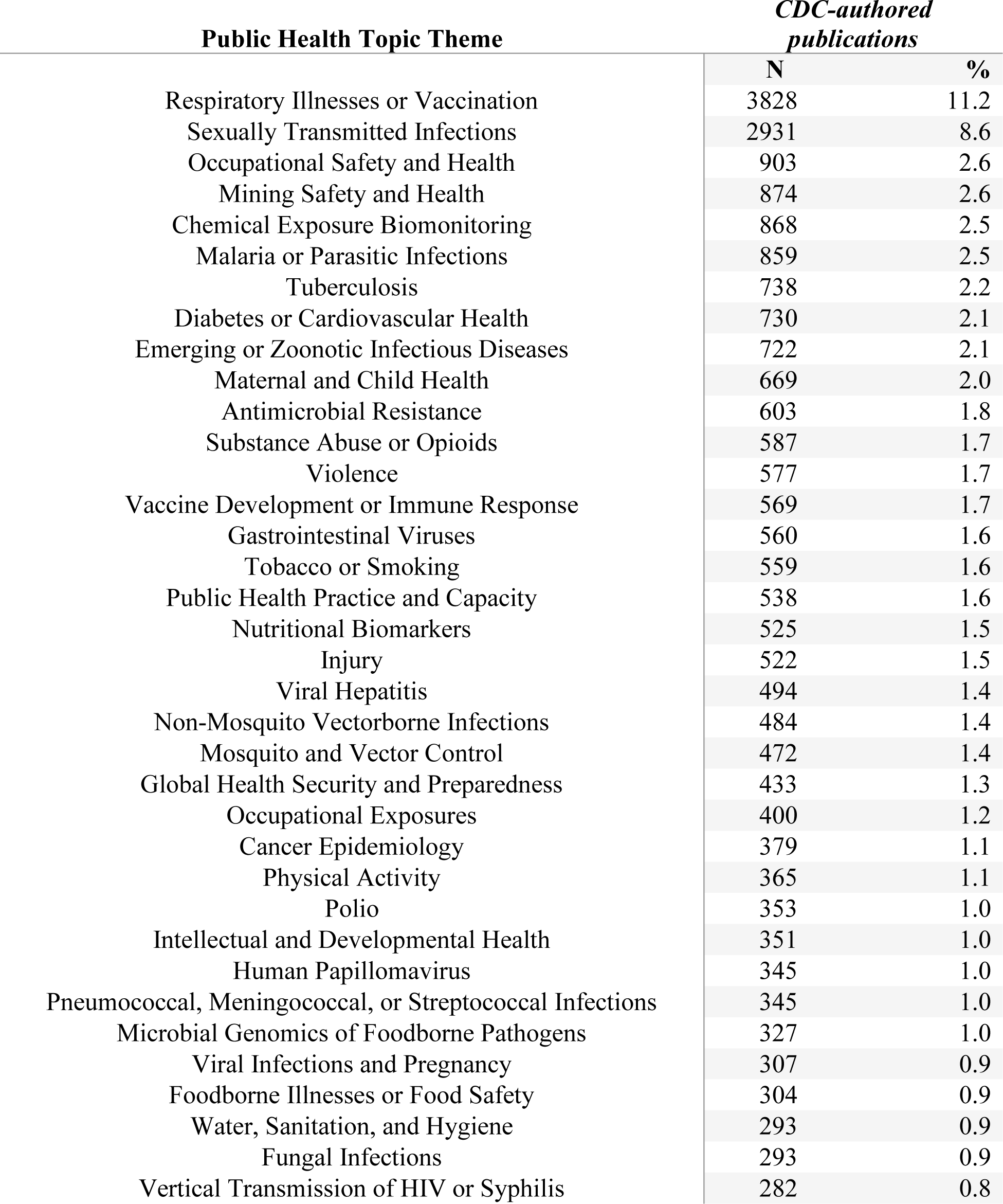
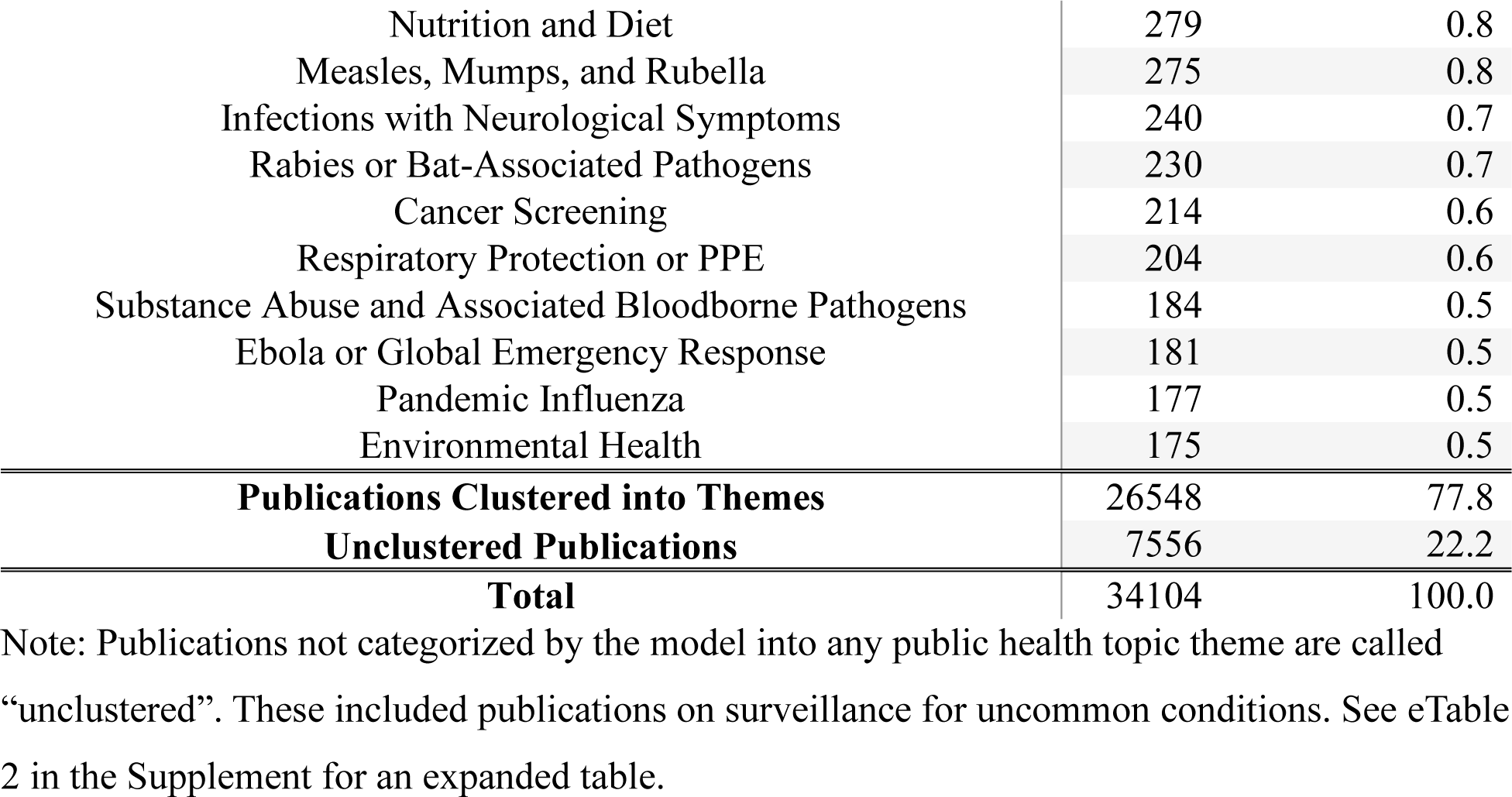
Number of CDC-authored publications by 46 public health topic themes identified by a large language model, 2014-2023.

**Figure 2.**
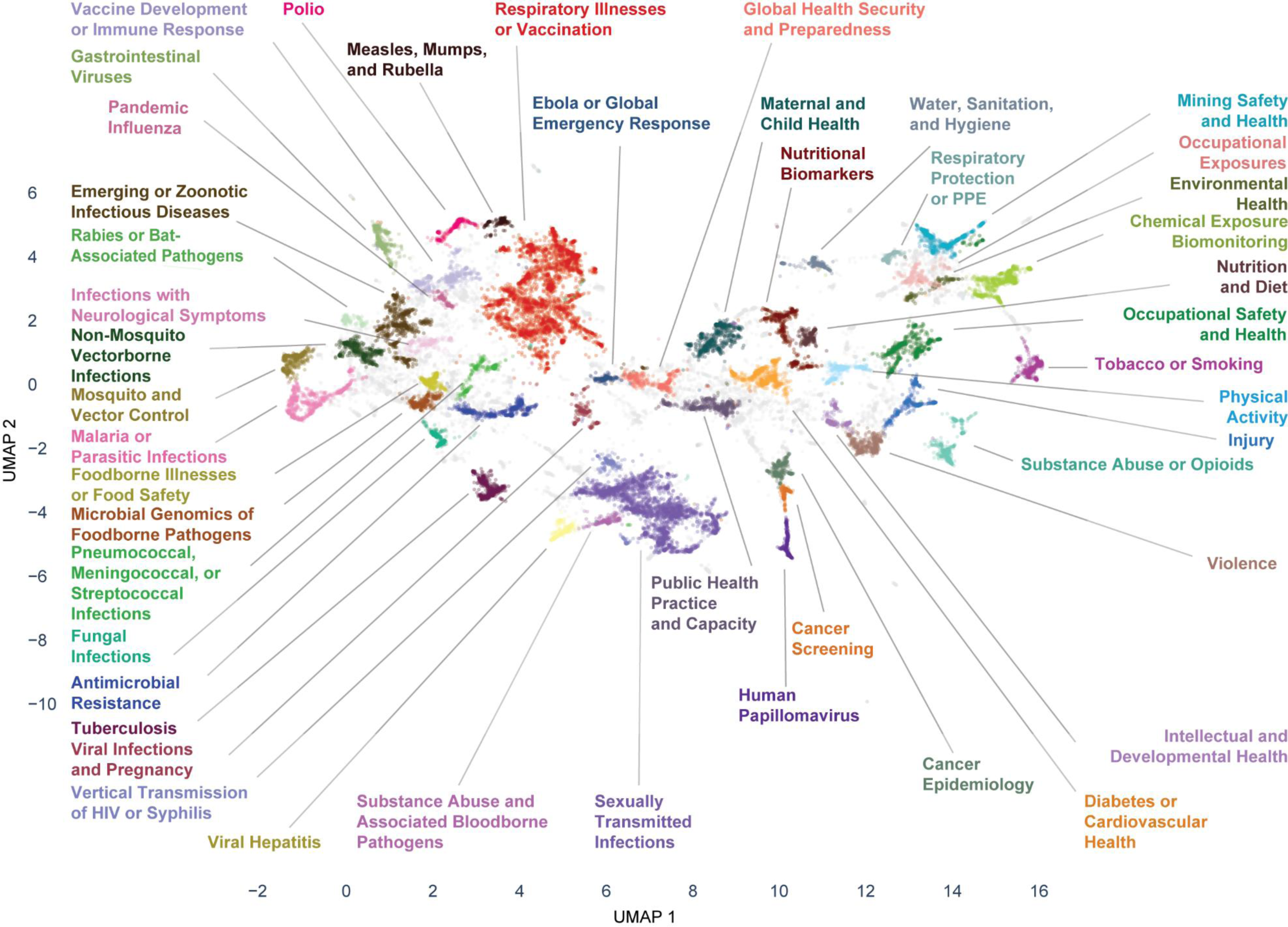
Topic map of CDC-authored publications by 46 public health topic themes identified by a large language model, 2014-2023. The 46 public health topic themes were identified by the optimized BERTopic topic modeling pipeline. To visualize relationships between topic themes, the publication embeddings were reduced in dimensionality using UMAP to two dimensions using the same nearest neighbors and minimum distance as the full pipeline. Similar papers are located closer together in space, while dissimilar papers are further apart. Each paper is represented as a point, and each public health topic theme has a unique color. Final labels for the public health topic themes were determined by reviewers. Gray points are papers that were unclustered, or not categorized into a public health topic theme by the model. Each point has an opacity of 0.25 to convey the density of points. Alt Text: Scatterplot illustrating a topic map of 46 public health topic themes. Each point in the scatterplot is a publication, and its color categorizes it as a member of one of the topic themes or unclustered.

Three public health topic themes directly related to agency-wide responses during the time period: Ebola or global emergency response (2014 Ebola Response), viral infections and pregnancy (2016 Zika Response), and respiratory illnesses or vaccination (2020 COVID-19 Response).^10,11^ These topic themes had keywords that reflected these response activities (eTable 1 in the Supplement). Notably, these topic themes also had the most publications in either the year of or the year after initiation of response activities (Figure 3).

**Figure 3.**
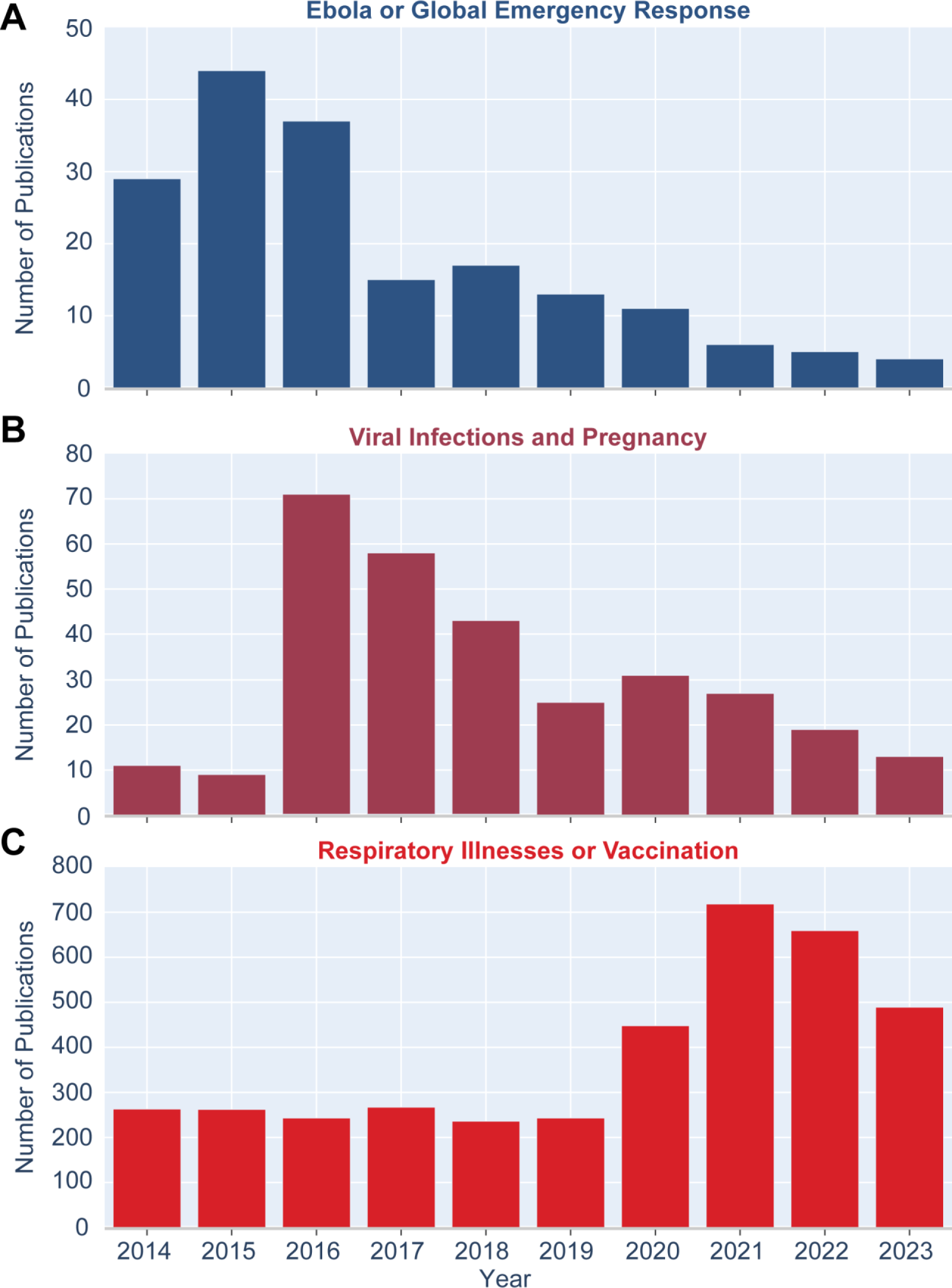
Number of publications in public health emergency response topic themes including “Ebola or Global Emergency Response” (A), “Viral Infections and Pregnancy” (B), and “Respiratory Illnesses or Vaccination” (C) – CDC-authored publications, 2014-2023. Alt Text: Bar charts labeled A to C. Bar charts show the number of publications by year.

CDC-authored publications in each public health topic theme had measurable academic, policy, and attention impact (eTable 2 in the Supplement). When ranking topic themes by three impact metrics--median academic citations (for publications 2014–2020), percent with policy citations (for publications 2014–2020), and median Altmetric Attention Score (for publications 2014– 2023)–we found that 13 different topics comprised the top 5 topics by each of the three impact metrics (Table 2). Two topics themes — fungal infections; substance abuse or opioids — were in the top 5 for two impact metrics. Of all topic themes, 28% were represented in this top 15, suggesting that diverse topics ranked highly on at least one impact metric. Conversely, no topic theme was ranked in the bottom quartile for all three metrics (eTable 2 in the Supplement). Some topic themes with a low ranking by one metric had a high ranking by another metric (e.g., fungal infections; mining safety and health), while other topic themes were more broadly impactful, ranking in the top 10 for all three impact metrics (e.g., respiratory illnesses or vaccination; substance abuse or opioids; intellectual and developmental health).

**Table 2.** Top 5 public health topic themes with the highest median academic citations, % with any policy citations, and median Altmetric Attention Score in CDC-authored publications, 2014-2023

| Rank | Median Number of Academic Citations* | Percent of Publications with Any Policy Citations* | Median Altmetric Attention Score ** |
| --- | --- | --- | --- |
| 1 | Fungal Infections*<br>(36.5) | Mining Safety and Health<br>(92.5%) | Substance Abuse or Opioids*<br>(14.0) |
| 2 | Substance Abuse or Opioids*<br>(29.5) | Occupational Safety and Health<br>(82.3%) | Viral Infections and Pregnancy<br>(11.0) |
| 3 | Chemical Exposure Biomonitoring<br>(29.0) | Respiratory Protection or PPE<br>(79.9%) | Respiratory Illnesses or Vaccination<br>(10.0) |
| 4 | Violence<br>(27.0) | Occupational Exposures<br>(79.7%) | Fungal Infections*<br>(10.0) |
| 5 | Intellectual and Developmental Health<br>(26.0) | Tuberculosis<br>(68.4%) | Foodborne Illnesses or Food Safety<br>(8.0) |

\* among 24,208 CDC-authored publications, 2014-2020
\*\* among 34,104 CDC-authored publications, 2014-2023
Note: The numeric values for these metrics are displayed in parentheses for each public health topic theme. Topic themes that appear more than once in this table are marked with an asterisk. For a complete list of these measures for all 46 public health topic themes, please see eTable 2 in the Supplement.

## DISCUSSION

From 2014-2023, CDC’s scientific publications had measurable academic, policy, and attention impact across at least 46 public health topic themes. These themes spanned infectious disease, non-infectious disease, and public health preparedness and response. Nearly all publications received academic citations, most received attention in news and social media, and more than half were cited in policy and clinical guidance. No public health topic theme studied by CDC authors ranked in the bottom quartile for all three impact metrics.

More than half of CDC-authored publications were cited in public health policy or clinical guidance. In contrast, a previous 2021 study found that approximately 6% of scientific publications receive policy citations.^19^ This same study showed that for all public health publications from 2008-2016 indexed in Scopus, 28% received policy citations. Another study in 2023 showed that 13% of publications generated through a large, federally funded program received policy citations.^21^ While these comparisons are affected by the date ranges of publications, the access date, and the database sources, they provide a baseline against which we can demonstrate that CDC-authored publications have high policy impact.

CDC-authored publications had higher attention in news and social media than baseline scientific publications and comparable impact to public health publications. From 2014-2023, approximately 69% of all publications indexed in Altmetric received attention (accessed November 27, 2024), compared to 84% of all CDC-authored publications (accessed July 2, 2024). However, 87% of Altmetric publications in this time period in the subject areas that were the top 3 most frequent first tagged in CDC-authored publications (i.e. Biomedical and Clinical Science, Health Sciences, and Biological Sciences) received attention (accessed November 27, 2024), which is more comparable to CDC’s metric. This could highlight an opportunity for improving communication strategies to ensure work reaches appropriate audiences.

A key strength of our study is consideration of three different types of impact metrics, which provide a holistic overview of how papers achieved scientific impact. Approaches that incorporate multiple types of metrics have been championed over merely analyzing academic citations or journal impact factors.^17,22,23^ Traditional bibliometric indicators are influenced by a variety of factors including the article’s field of study, and efforts have been made to normalize for these field-specific trends in academic citations, notably through the Relative Citation Ratio.^7,18,24^ Field-specific trends for attention metrics and policy citations have also been demonstrated.^19,25^ Even without normalizing for public health subdisciplines, we find that no topic theme in our model ranked in the bottom quartile of themes for all three metrics, demonstrating that each topic theme had non-negligible impact in either media attention, academia, or policy.

The public health topic themes in our model reflect CDC’s past and current strategic science priorities. All major CDC centers are represented by the topic themes.^26^ Our model produced topic themes related to several agency-wide public health responses, and CDC scientists may have shifted priorities to ensure high volume of publications related to these responses (Figure 3). Notably, these themes emerged from the model without explicitly instructing it to produce clusters related to the agency’s known priorities.

Topic models can be used to evaluate shifts in important scientific topics and to identify opportunities to prioritize topics moving forward, whether that means initiating research into emerging topics or finding new strategies to increase media attention or policy translation for specific topics. For example, previous work combining topic tagging with bibliometrics demonstrated the scale and impact of CDC’s COVID-19 publications.^6^

The use of topic modeling in bibliometrics has evolved from earlier applications that used word co-occurrences and Latent Dirichlet Allocation (LDA)^27–29^ towards large language models.^12,30,31^ One of the strengths of topic modeling for these applications is the ability to characterize large publication portfolios quickly.^27–30^ Developing reliable search strategies for the entire range of topics addressed by authors at any large research institution such as CDC would likely be time and resource prohibitive. Approaches that rely on subject matter experts to categorize papers or develop systematic search strategies can also introduce bias. Other field classification systems can be too general, or poorly reflect emerging topics.^28^ Alternatively, topic modeling as performed in this study can be considered “unsupervised,” meaning the model was not constrained to identify specific topics. As a result, the model may be less prone to operator bias and more acute to topic themes and similarities between papers that a set of systematic search strategies could miss.

Our analysis is subject to several limitations. First, our metrics are not strictly independent of each other. For example, the number of policy mentions contributes (though with lower weighting than news articles) to the Altmetric Attention Score.^32^ Similarly, some studies have shown correlations between attention metrics, academic citations, and policy citations.^20,21^ Second, our analysis may not capture all CDC-authored publications (e.g., any not listed in Science Clips), or their full impact (e.g., from any not indexed in Altmetric or BMJ Impact Analytics), thus underestimating the true impact of CDC science. We conducted quality checks using internal data (not shown), which indicated that the version of Science Clips used in this study captures >95% of CDC-authored publications during the studied time period. Missingness of Altmetric data was very small (approx. 0.3% of publications) but had an outsized effect on small clusters (e.g. 10% of environmental health publications, 17/175, were missing Altmetric data). Third, a publication’s mentions in media and its academic citations do not always indicate public health impact, as some studies have previously explored.^17,20,33,34^ Fourth, our topic model was influenced by our parameter choices. The dimensionality reduction technique used in BERTopic, UMAP, is stochastic, so results vary not only from parameter choices, but also based on the machine used to optimize the model and the random state chosen.^14^ We set the random state for reproducibility, but stochasticity could have produced a model with a different number of topic themes or structure. We mitigated this limitation through human review of our model, which demonstrated that the categories generated by our model were reasonable. Searching across random states to ensure that results are robust to this parameter would improve the optimization strategy. Fifth, not all publications in our list were classified into a topic theme, so it is possible that we missed publications that could have contributed to overall impact of certain topic themes. Finally, some topic themes in our model, especially the largest ones, included outliers that might have been removed using a systematic search strategy.

## CONCLUSIONS

We evaluated impacts of CDC-authored publications from 2014-2023 by topic theme using multiple objective measures. Topic modeling was crucial for identifying topic themes in a diverse publication portfolio. Multiple impact metrics allowed us to evaluate how topic areas disseminated evidence and affected public health policy and clinical guidance. Our methods can be applied to other organizations seeking to evaluate impacts of large, diverse publication portfolios. These metrics can inform further assessment of specific topic or programmatic areas with an aim to strengthen science, policy, and communications partnerships to increase science translation that can lead to improving health and saving lives. Providing this overview of the content and impact of CDC science promotes transparency into the activities of the agency with the public and partners.

## Data Availability

All code and data produced are available online in a GitHub repository.

https://github.com/cdcai/analysis-bertopic-cdc-publications

## ACKNOWLEDGMENTS

The authors would like to acknowledge the CDC Office of Science leadership and Knowledge Management teams for their support of this project. They would also like to acknowledge current and past members of the CDC Publications Portfolio Work Group who contributed feedback on this project.

Portions of this work were presented at the American Medical Informatics Association (AMIA) 2024 Annual Symposium in San Francisco, CA.

## CODE AND DATA AVAILABILITY

Code, example notebooks, and data are available through a GitHub repository: cdcai/analysis-bertopic-cdc-publications.

Science Clips is freely available as a public-facing database: https://phgkb.cdc.gov/PHGKB/cdcSCStartPage.action.

## Disclaimer

The findings and conclusions in this report are those of the authors and do not necessarily represent the official position of the Centers for Disease Control and Prevention.

## SUPPLEMENTARY FILES

- **eText.** BERTopic Optimization Strategy – Coverage, Relative Validity Score, and Evenness.
- **eFigure 1.** Number of academic citations, number of Altmetric policy citations, and number of BMJ Impact Analytics policy citations by publication year – CDC-authored publications, 2014-2023.
- **eTable 1.** Notes from Topic Model Review by CDC Subject Matter Experts.
- **eTable 2.** Number and percent of publications, median number of academic citations, % with any policy citations, median Altmetric Attention Score, academic metric rank, policy metric rank, and attention metric rank, for 46 public health topic themes identified by a large language model – CDC-authored publications, 2014-2023.

