## Supplementary Material for "From Research to Impact: Assessing a Decade of CDC’s Public Health Science by Topic Area, 2014-2023"

**Supplementary Text.** *BERTopic Optimization Strategy – Coverage, Relative Validity Score, and Evenness.*

To create a topic model for CDC-authored publications, we used a grid search optimization strategy to find a BERTopic modeling pipeline that produced a well-behaved model.[1] A well-behaved model would have clusters that are interpretable and distinguishable by human subject matter experts, with few outlier papers within clusters. We found that three measures were useful for optimization: *coverage*, the *relative validity score*[2], and *evenness*. We weighted these three measures equally to determine the *overall score* of a model.

*Coverage* was defined as the percent of publications that were placed into a cluster (Eq. 1):

$$Coverage = \frac{N_{total} - N_{unclustered}}{N_{total}}$$

where $N_{total}$ is the total number of publications and $N_{unclustered}$ is the number of unclustered publications. $N_{total} - N_{unclustered}$ would equal the number of clustered publications.

The *relative validity score* is a fast calculation of the *Density Based Cluster Validity (DBCV) score*[2] available in the HDBSCAN clustering library[*]. It is a relative measure of the goodness of clustering compared to other models, and it is specifically designed for use with clustering algorithms like HDBSCAN[3].

*Evenness* is a measure we implemented after observing that some models with high coverage or relative validity scores created large clusters that did not distinguish well between public health topic themes. *Evenness* was defined as (Eq. 2):

$$Evenness = 1 - \frac{C_{max} - C_{min}}{N_{total} - 2}$$

where $C_{max}$ is the number of publications in the largest cluster, $C_{min}$ is the number of publications in the smallest cluster, and $N_{total}$ is the total number of publications. *Evenness* is 0 when every publication is clustered, $C_{max} = N_{total} - 1$, and $C_{min} = 1$. *Evenness* is 1 when $C_{max} = C_{min}$. There may be other methods of calculating a measure that scores the model based on how similar

[*] https://hdbscan.readthedocs.io/en/latest/index.html

clusters are in size. Here, a higher scoring model will have similarly sized largest and smallest clusters.

The *overall score* for a model was calculated as (Eq. 3):

$$Overall\ Score = \frac{1}{3} \times Coverage + \frac{1}{3} \times Relative\ Validity\ Score + \frac{1}{3} \times Evenness$$

All three measures were equally weighted in the overall score. These weightings could be altered to favor one measure over another.

Our grid search tested 3,600 unique combinations of UMAP[4] and HDSBCAN parameters. Each possible combination of the following values was tested:

| **Number of Neighbors** | **Number of Dimensions** | **Minimum Distance** | **Minimum Cluster Size** | **Minimum Samples** |
|---|---|---|---|---|
| 15 | 25 | 0.00 | 25 | 10 |
| 25 | 50 | 0.25 | 50 | 20 |
| 50 | | 0.50 | 75 | 30 |
| | | 0.75 | 100 | 40 |
| | | 1.00 | 125 | 50 |
| | | | 150 | 60 |
| | | | 175 | 70 |
| | | | 200 | 80 |
| | | | 225 | 90 |
| | | | 250 | 100 |
| | | | | 110 |
| | | | | 120 |

Other UMAP parameters that could be included in a grid search include *metric* (for computing distances) and *random state* (for setting initial random state of clusters), among others. Other HDBSCAN parameters that could be included in a grid search include *metric* (for computing distances) and *cluster selection method* (which can affect size of clusters), among others. For *metric* in both UMAP and HDBSCAN and *cluster selection method*, we used the default parameters recommended in BERTopic documentation. We set *random state* to ensure no stochasticity in our grid search.

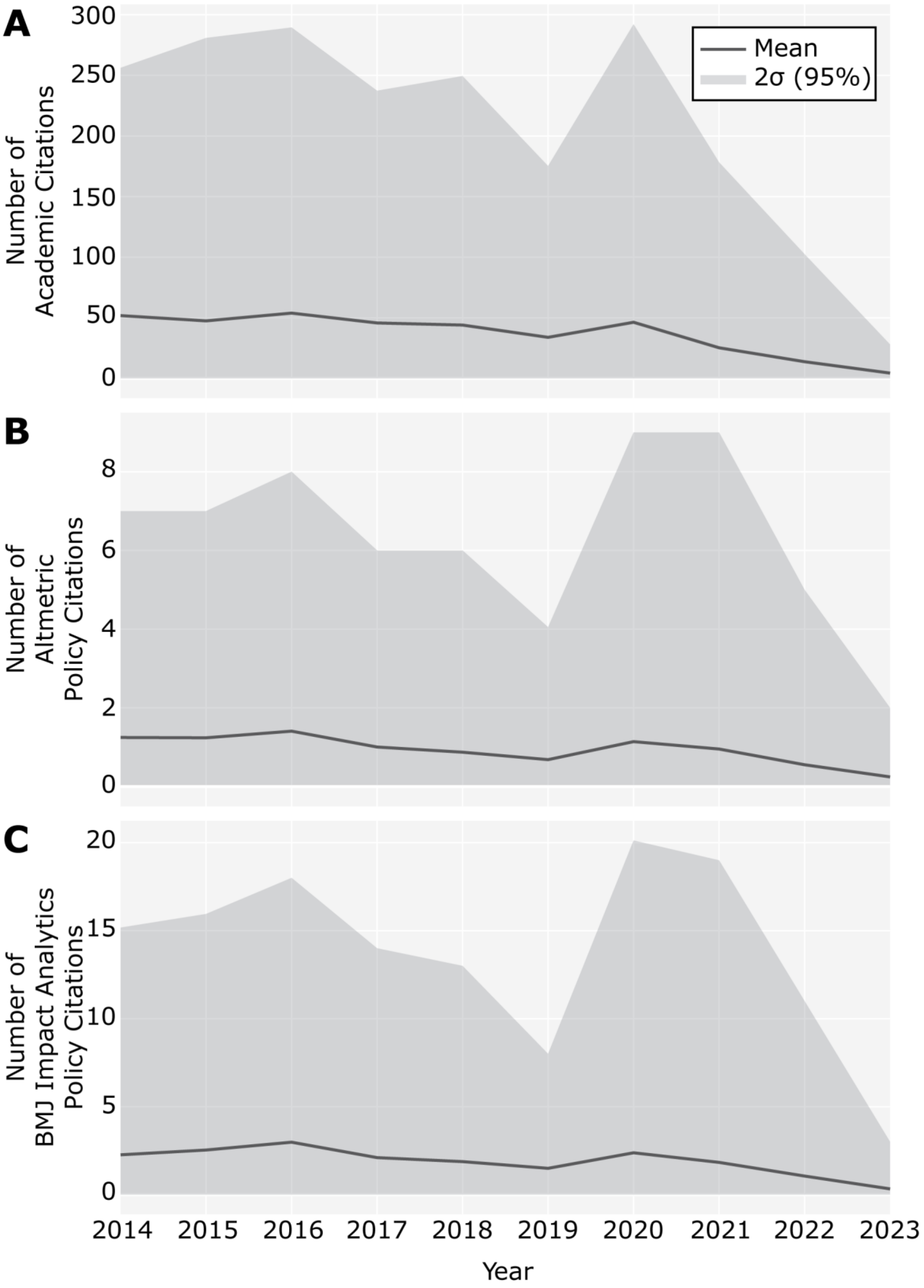

A
B
C
Mean
2σ (95%)
Number of Academic Citations
Number of Altmetric Policy Citations
Number of BMJ Impact Analytics Policy Citations
300
250
200
150
100
50
0
8
6
4
2
0
20
15
10
5
0
2014
2015
2016
2017
2018
2019
2020
2021
2022
2023
Year


55

**Supplementary Figure 1.** *Number of academic citations, number of Altmetric policy citations, and number of BMJ Impact Analytics policy citations by publication year – CDC-authored Publications, 2014-2023*. The mean (gray line) ± approximately two standard deviations, 2σ (dark gray area; from the 2.5$^{th}$ to the 97.5$^{th}$ percentile) is shown for the number of academic citations (A), the number of Altmetric policy citations (B), and the number of BMJ Impact Analytics policy citations (C) for CDC-authored journal articles by publication year, as of July 2, 2024.

Alt Text: Graphs labeled A to C. Graphs and data show the mean and approximately two standard deviations of the number of academic citations, number of Altmetric policy citations, and number of BMJ Impact Analytics policy citations by publication year.

**Supplementary Table 1.** *Notes from Topic Model Review by CDC Subject Matter Experts.*

| Topic Label | Topic Model Keywords | Reviewer A | Reviewer B | Reviewer C | Reviewer D | Stage 1 Label | Stage 2 Notes | Final Label |
|---|---|---|---|---|---|---|---|---|
| 1 | ['cov infection', 'vaccine effectiveness', 'influenza vaccine', 'vaccination coverage', 'influenza vaccination', 'hospitalizations', '19 vaccination', 'vaccine', 'influenza', 'pneumonia'] | Vaccine Effectiveness | COVID-19 and Influenza | Vaccine - Preventable Epidemic Prone Infections | Vaccine safety/effectivness, COVID, Influenza, Respiratory illnesses, Vaccination/imunization programs/uptake,coverage | Respiratory Illnesses and Vaccination | Stage 2 reviewers noted that this cluster contains vaccination studies for viruses that are non-respiratory in nature. This is the largest cluster and can be non-specific. Stage 2 reviewers requested clarifying any label where "and" occurs. Some of these can be better reflected as an "or" to indicate that the paper doesn't have to be about both sub-topics. | Respiratory Illnesses or Vaccination |
| 2 | ['antiretroviral therapy', 'chlamydia', 'sexually transmitted', 'condom', 'hiv prevention', 'gonorrhea', 'hiv infection', 'viral load', 'sex men', 'hiv care'] | Sexually Transmitted Diseases | Sexually Transmitted Diseases | Sexually-transmitted Infections--Trends, Detection, Prevention, Treatment | Sexually transmitted diseases/infections, HIV, sexual/reproductive health education, contraceptive use, antiretrovirals | Sexually Transmitted Infections | | Sexually Transmitted Infections |
| 3 | ['occupational health', 'workplace', 'occupational safety', 'workforce', 'workers', 'institute occupational', 'employees', 'safety health', 'workers compensation', 'occupational'] | Occupational safety and health (combine 2, 3, and 23) | Occupational Health and Safety | Occupational Health--Injury, Risk & Protective Factors, Disparities | Worker safety, occupational health/safety /injuries, Sleep, Worker wellness | Occupational Safety and Health | Stage 2 reviewers categorized some papers in this cluster as Occupational Exposures. Stage 2 reviewers requested clarifying any label where "and" occurs. Some of these can be better reflected as an "or" to indicate that the paper doesn't have to be about both sub-topics. | Occupational Safety and Health |

| | | | | | | | | |
|---|---|---|---|---|---|---|---|---|
| 4 | ['respirable dust', 'aerosols', 'sampling', 'respirable', 'dust', 'occupational safety', 'fume', 'aerosol', 'rock', 'particles'] | Occupational safety and health (combine rank 2, 3, and 23) | Respiratory Health | Occupational Risk Characterization & Mitigation--Innovation, Measures | Mining safety, mining structure, mining inhalation exposures (dust, fungal), exposures and lung tissue, nanomaterials exposure to lung tissue/mice/ rat models | Mining Safety and Health | Stage 2 reviewers categorized some papers in this cluster as Water Safety and Hand Hygiene. Stage 2 reviewers requested clarifying any label where "and" occurs. Some of these can be better reflected as an "or" to indicate that the paper doesn't have to be about both sub-topics. | Mining Safety and Health |
| 5 | ['phthalates', 'urinary concentrations', 'phthalate', 'urine samples', 'bisphenol', 'exposures', 'pfas concentrations', 'chemicals', 'environmental', 'urine'] | Environmental monitoring (combine 4 with 45) | Chemical Exposures | Environmental Hazards--Exposure, Biomarkers, PFAS, Outcomes | Biomonitoring/biomarkers of environmental/chemical exposures or smoking/cannabis exposure, paternal/maternal exposures | Chemical Exposure Monitoring | reviewers overlapped this topic with Occupational Exposures. One distinction of this cluster is that most papers involve biomonitoring in people through urinary concentrations, blood serum, plasma, etc. It is focused on the methods used. | Chemical Exposure Biomonitoring |
| 6 | ['malaria cases', 'malaria transmission', 'malaria control', 'antimalarial', 'malaria pregnancy', 'malaria', 'schistosomiasis', 'plasmodium falciparum', 'falciparum', 'rapid diagnostic'] | Mosquito-borne diseases (combine 5, 21 and 31) | Malaria | Parasitic Infections--Global Burden, Detection, Treatment | Vector borne diseases, parasitic infections, malaria, chagas, Schistosomiasis, various worm parasites | Malaria and Parasitic Infections | Stage 2 reviewers requested clarifying any label where "and" occurs. Some of these can be better reflected as an "or" to indicate that the paper doesn't have to be about both sub-topics. | Malaria or Parasitic Infections |
| 7 | ['resistant tuberculosis', 'tuberculosis infection', 'tuberculosis tb', 'latent tuberculosis', 'tb | Tuberculosis (TB) | Tuberculosis | Tuberculosis--Global Surveillance, Drug Susceptibility, Testing | Tuberculosis, screening for TB | Tuberculosis | | Tuberculosis |

| | | | | | | | | |
|---|---|---|---|---|---|---|---|---|
| | infection', 'rifapentine', 'mycobacterium tuberculosis', 'multidrug resistant', 'tuberculosis', 'drug resistant'] | | | | | | | |
| 8 | ['diabetes prevention', 'adults diabetes', 'type diabetes', 'diagnosed diabetes', 'diabetes', 'prediabetes', 'cardiovascular disease', 'kidney disease', 'heart disease', 'care'] | Diabetes | Diabetes and Cardiovascular Health | Chronic Diseases--Trends, Disparities, Costs | cardiovascular disease, kidney disease, diabetes, heart attack, stroke, hypertension, NHANES | Diabetes and Cardiovascular Health | Stage 2 reviewers requested clarifying any label where "and" occurs. Some of these can be better reflected as an "or" to indicate that the paper doesn't have to be about both sub-topics. | Diabetes or Cardiovascular Health |
| 9 | ['dengue virus', 'leptospirosis', 'leptospira', 'chikungunya virus', 'arboviral', 'dengue', 'monkeypox virus', 'yellow fever', 'encephalitis', 'febrile illness'] | Tropical diseases | Emerging Infectious Diseases | Zoonotic Diseases--Environment, Ecology, Transmission | emerging infectious diseases, neglected infectious diseases, viral taxonomy, epidemiological characteristics or transmission of rare infectious diseases, especially viral | Emerging Infectious Diseases | Stage 2 reviewers categorized papers in this cluster as Immune Response and Vaccine Development or Mosquito and Vector Control, among others. Stage 2 reviewers requested the topic label be changed to reflect the major CDC center (National Center for Emerging and Zoonotic Infectious Diseases). | Emerging or Zoonotic Infectious Diseases |
| 10 | ['birth defects', 'stillbirths', 'obstetric', 'preterm birth', 'live births', 'maternal mortality', 'national birth', 'infant mortality', 'gestational age', 'congenital heart'] | Birth defects | Maternal and Infant Health | Maternal & Infant Health--Congenital Disorders, Mortality, Disparities | birth/congenital defects, perinatal health care, maternal/infant mortality, pregnancy risks, safe sleep | Maternal and Infant Health | Stage 2 reviewers suggested changing the label of this category to "Maternal and Child Health". | Maternal and Child Health |

| | | | | | | | | |
|---|---|---|---|---|---|---|---|---|
| 11 | ['antibiotic stewardship', 'antimicrobial stewardship', 'antibiotic prescribing', 'antibiotic use', 'antibiotic resistance', 'carbapenem resistant', 'acute care', 'associated infections', 'antimicrobial resistance', 'carbapenem'] | Antimicrobial resistance | Antibiotic Stewardship | Antimicrobial Resistance--Detection, impact, Stewardshiup | Antimicrobial use/stewardship/prescribing, antibiotic resistance, healthcare associated infections | Antimicrobial Resistance | Stage 2 reviewers categorized one paper as Maternal and Infant Health, and another as Pneumococcal, Meningococcal, and Streptococcal Infections. | Antimicrobial Resistance |
| 12 | ['opioid overdose', 'prescription opioids', 'drug overdose', 'prescription opioid', 'opioid use', 'overdoses', 'opioid prescribing', 'opioid prescriptions', 'opioids', 'overdose deaths'] | Opioid overdose | Overdose Prevention | Opioids and Substance Abuse--Trends, Prevention | Oversdoses, Substance abuse, alcohol use, opioids, neonatal exposure to illicit substances, poisoning/exposure to drugs of children | Substance Abuse and Opioids | Stage 2 reviewers requested clarifying any label where "and" occurs. Some of these can be better reflected as an "or" to indicate that the paper doesn't have to be about both sub-topics. | Substance Abuse or Opioids |
| 13 | ['violence children', 'youth violence', 'partner violence', 'sexual violence', 'violence prevention', 'physical violence', 'violence victimization', 'dating violence', 'violence perpetration', 'youth risk'] | Domestic violence | Violence Prevention in Youth | Violence--Risk & Protective Factors, Childhood Experiences | mostly Adverse childhood experiences, youth/child violence/suicide, gender based/sexual violence, some viedo game interventions for violence prevention | Violence | Stage 2 reviewers sometimes categorized papers in this cluster as Injury or Maternal and Infant Health. | Violence |

| | | | | | | | | |
|---|---|---|---|---|---|---|---|---|
| 14 | ['influenza vaccine', 'influenza vaccines', 'vaccine', 'antibody responses', 'influenza virus', 'immunogenicity', 'influenza viruses', 'immune responses', 'immune response', 'antibody titers'] | Vaccine development | Immune Response Studies | Viral Infections--Vaccine & Infection Induced Immune Responses | Immunogenicity/pathogenicity/immune response to antivirals, viruses, and vaccination | Immune Response and Vaccine Development | Stage 2 reviewers disagreed with the model and categorized some papers in this cluster as Respiratory Illnesses and Vaccination or Emerging Infectious Diseases. Stage 2 reviewers requested that vaccine development be listed first since it is more public health focused. Stage 2 reviewers requested clarifying any label where "and" occurs. Some of these can be better reflected as an "or" to indicate that the paper doesn't have to be about both sub-topics. | Vaccine Development or Immune Response |
| 15 | ['rotavirus vaccine', 'rotavirus vaccines', 'rotavirus gastroenteritis', 'rotavirus vaccination', 'rotavirus infection', 'rotaviruses', 'rotavirus', 'vaccine effectiveness', 'vaccine introduction', 'gastroenteritis age'] | Rotavirus vaccines | Rotavirus | Gastrointestinal Viruses--Prevention, Burden | Diarrheal illness (viral cause), gastroenteritis, rotavirus, Caliciviridae (norovirus, sapovirus, etc), enteroviruses (polio, etc) | Gastrointestinal Viruses | | Gastrointestinal Viruses |

| | | | | | | | | |
|---|---|---|---|---|---|---|---|---|
| 16 | ['tobacco use', 'tobacco products', 'adult tobacco', 'tobacco product', 'cigarette use', 'youth tobacco', 'tobacco control', 'use tobacco', 'tobacco survey', 'marijuana use'] | Tobacco use | Tobacco Use | Tobacco--Cessation, Usage Trends, Policy | Smoking, tobacco, nicotine, vaping/e-cigarettes, marijuana | Tobacco and Smoking | Stage 2 reviewers requested clarifying any label where "and" occurs. Some of these can be better reflected as an "or" to indicate that the paper doesn't have to be about both sub-topics. | Tobacco or Smoking |
| 17 | ['health workforce', 'health departments', 'public health', 'epidemiology training', 'health care', 'health equity', 'accreditation', 'workforce', 'interventions', 'practice'] | Workforce health | Public Health Infrastructure | Public Health Workforce, Community, Partnerships, Accountability | public health workforce/research/legal/evaluation capacity, priorities/policy/strategy, data standards, collaborations/partnerships, metaresearch/bibliometrics, scientific writing, diversity/disparities, social determinants of health | Public Health Practice and Capacity | Stage 2 reviewers noted that many papers could be considered Public Health Practice and Capacity even if there was a cluster for which they had a better fit. Stage 2 reviewers requested clarifying any label where "and" occurs. Some of these can be better reflected as an "or" to indicate that the paper doesn't have to be about both sub-topics. | Public Health Practice and Capacity |
| 18 | ['iron deficiency', 'folate concentrations', 'vitamin b12', 'micronutrient', 'folic acid', 'vitamin 12', 'nutrition', 'folate', 'nutritional', 'vitamin'] | Anemia | Micronutrients | Bodily Health--Biomarkers, Nutrition, BMI | nutrition, obesity, human blood/serum/plasma, anemia, blood disorders, macro/micronutrients, vitamins | Nutritional Biomarkers | | Nutritional Biomarkers |
| 19 | ['injury prevention', 'violent death', 'injuries', 'fatalities', 'injury', 'homicide', 'homicides', 'brain | Violence Prevention | Violence and Injury Prevention | Injury--Impacts, Trends, Risk Factors | injuries and deaths, traumatic brain injuires, suicides, ER visits, motor | Injury | Stage 2 reviewers sometimes categorized papers in this cluster as Violence. | Injury |

| | | | | | | | | |
|---|---|---|---|---|---|---|---|---|
| | injury', 'violence', 'trauma'] | | | | vehicle/firearm/sports related injury/death | | | |
| 20 | ['chronic hepatitis', 'hepatitis vaccine', 'viral hepatitis', 'hepatitis virus', 'hbv infection', 'hepatitis vaccination', 'hcv infection', 'hepatitis surface', 'hepatitis', 'liver disease'] | Viral hepatitis | Hepatitis | Hepatitis--Epidemiology, Prevention, Treatment, Disparities | hepatitis, liver disease/cancer, Cirrhosis, kidney transplant | Viral Hepatitis | | Viral Hepatitis |
| 21 | ['tick borne', 'ticks collected', 'tick bite', 'ticks', 'tickborne', 'pestis', 'spirochetes', 'burgdorferi', 'borrelia burgdorferi', 'burgdorferi sensu'] | Tick borne diseases | Tickborne Illnesses | Vector-borne Infections | ectoparasitic arthropods, tickborne illnesses, fleaborne illnesses, rodent transmitted illnesses, Vector borne diseases, parasitic diseases, | Non-Mosquito Vectorborne Infections | | Non-Mosquito Vectorborne Infections |
| 22 | ['aedes aegypti', 'insecticide resistance', 'anopheles gambiae', 'aegypti', 'ae aegypti', 'malaria vectors', 'mosquito control', 'malaria vector', 'insecticidal nets', 'pyrethroid resistance'] | Mosquito-borne diseases (combine 5 with 21) OR combine with Zika virus (rank 31) as well | Mosquito Control | Mosquito-borne Infections & Vector Control | vector control/management, insecticides, mosquito control, mosquitos | Mosquito and Vector Control | Stage 2 reviewers sometimes categorized papers in this cluster as Malaria and other Parasitic Infections. | Mosquito and Vector Control |

| | | | | | | | | |
|---|---|---|---|---|---|---|---|---|
| 23 | ['emergency preparedness', 'health preparedness', 'emergency response', 'health emergency', 'preparedness response', 'preparedness', 'public health', 'emergency management', 'emergency', 'disasters'] | Emergency preparedness | Readiness and Response | Global Public Health Security | preparedness, emergency/disaster/surveillance response, global health security, public health threats, global health community | Global Health Security | Stage 2 reviewers requested that this label reflect papers in the cluster on emergency preparedness. | Global Health Security and Preparedness |
| 24 | ['asbestos', 'occupational exposure', 'coal workers', 'exposures', 'cancer mortality', 'radon', 'lung cancer', 'exposure', 'cohort', 'respiratory health'] | Occupational safety and health (combine topics 2, 3, and 23) | Occupational Exposures | Environmental Contaminants & Hazardous Occupational Exposures | occupational exposures, air quality/exposure, environmental health, radiation/radioactive exposure, environmental fungal exposure/allergens, pollution exposure | Occupational Exposures | Stage 2 reviewers disagreed with the model and categorized some papers in this cluster as Environmental Health and Respiratory Protection. Changes to the Respiratory Protection topic name may help distinguish categories. | Occupational Exposures |
| 25 | ['cancer registries', 'cancer incidence', 'cancer prevention', 'cancer registry', 'national cancer', 'cancer screening', 'cancer mortality', 'lung cancer', 'cancer patients', 'comprehensive cancer'] | Cancer prevention | Cancer Prevention | Cancer--Surveillance, Survivorship Trends, Disparities | cancer | Cancer Epidemiology | Stage 2 reviewers noted some overlap with Cancer Screening cluster. | Cancer Epidemiology |
| 26 | ['physical activity', 'intervention', 'interventions', 'activity guidelines', 'exercise', 'public health', 'aerobic | Workplace wellness | Physical Activity | Physical Activity & Health--Trends, Barriers, Policy | physical activity, transportation, built environment, arthritis, | Physical Activity | | Physical Activity |

| | | | | | | | | |
|---|---|---|---|---|---|---|---|---|
| | physical', 'inactivity', 'sedentary', 'older adults'] | | | | osteoporosis, bone density | | | |
| 27 | ['polio vaccine', 'poliovirus vaccine', 'poliovirus transmission', 'inactivated poliovirus', 'poliovirus type', 'derived polioviruses', 'derived poliovirus', 'poliovirus wpv', 'oral poliovirus', 'polio eradication'] | Poliovirus | Poliovirus | Polio--Global Eradiciation Campaign | polio, acute flaccid paralysis, enteric vaccination | Polio | | Polio |
| 28 | ['autism spectrum', 'developmental disabilities', 'children asd', 'autism developmental', 'children autism', 'spectrum disorder', 'children aged', 'child', 'autism', 'disorder asd'] | Autism | Developme ntal Disorders | Pediatric Health--Development al Screening & Detection of Disability | intellectual and development al disabilities, autism, ADHD, pediatric/ad olescent mental health surveillance, schools/acad emic acheivement and health interventions for development | Intellectual and Developmental Health | Stage 2 reviewers requested clarifying any label where "and" occurs. Some of these can be better reflected as an "or" to indicate that the paper doesn't have to be about both sub-topics. | Intellectual and Developmental Health |
| 29 | ['papillomavirus vaccination', 'human papillomavirus', 'papillomavirus vaccine', 'papillomavirus hpv', 'cervical cancer', 'papillomavirus', | Human Papillomavi rus | Human Papillomavi rus | Human Papillomavir us -- Vaccination, Cancer Prevention & Screening | HPV, cervical cancer, pelvic exams, cervical cancer screening | Human Papillomavirus | | Human Papillomavirus |

| | | | | | | | | |
|---|---|---|---|---|---|---|---|---|
| | 'hpv vaccination', 'hpv vaccine', 'cancer screening', 'hpv infection'] | | | | | | | |
| 30 | ['invasive pneumococcal', 'pneumococcal disease', 'valent pneumococcal', 'meningococcal disease', 'serogroup meningococcal', 'pneumococcal conjugate', 'pneumococcal', 'meningitidis', 'pneumococcal meningitis', 'meningococcal'] | Pneumococcal Meningitis | Pneumococcal and Meningococcal Disease | Pneumococcal, Meningococcal, & Streptococcal Infections | pneumonia, pneumococcal infections/illness/vaccination, group a strep, bacterial meningitis, meningococcal illnesses/vaccination | Pneumococcal, Meningococcal, and Streptococcal Infections | Stage 2 reviewers requested clarifying any label where "and" occurs. Some of these can be better reflected as an "or" to indicate that the paper doesn't have to be about both sub-topics. | Pneumococcal, Meningococcal, or Streptococcal Infections |
| 31 | ['salmonella enterica', 'antimicrobial resistance', 'vibrio cholerae', 'cholerae', 'escherichia coli', 'sequencing wgs', 'salmonella', 'enterica', 'pneumophila', 'sequence typing'] | Foodborne Pathogens | Foodborne Bacterial Pathogens | Microbial Genomes--Molecular Typing, Population Analysis, Tools | genomics, genomic sequencing/characterization, genotyping, biofilms, taxonomic classification, phylogenetics | Microbial Genomics of Foodborne Pathogens | Stage 2 reviewers categorized one paper as Respiratory Protection, since it involved PPE in food preparation. Modifications were made to the Respiratory Protection label. | Microbial Genomics of Foodborne Pathogens |
| 32 | ['infection pregnancy', 'zikv infection', 'zika virus', 'virus exposure', 'birth defects', 'pregnant women', 'congenital cytomegalovirus', 'zika syndrome', 'transmission zika', 'virus disease'] | Zika virus OR Mosquito-borne diseases (combine topics 5, 21, and 31) | Viral Infections in Pregnancy | Acute Infections in Pregnancy & Post-pregnancy Outcomes | viral infection during pregnancy and congenital abnormalities/birth defects, zika, cytomegalovirus | Viral Infections and Pregnancy | Stage 2 reviewers found it difficult to distinguish between this cluster and Congenital Infections. | Viral Infections and Pregnancy |

| | | | | | | | | |
|---|---|---|---|---|---|---|---|---|
| 33 | ['foodborne diseases', 'foodborne disease', 'foodborne illness', 'disease outbreaks', 'foodborne', 'food safety', 'illness outbreaks', 'outbreak strain', 'escherichia coli', 'coli o157'] | Foodborne Illnesses | Foodborne Disease | Foodborne Illness & Food Safety | foodborne illness/disease outbreaks, food safety, foodborne illness detection/surveillance, probiotics, salmonella, food allergy | Foodborne Illnesses and Food Safety | Stage 2 reviewers requested clarifying any label where "and" occurs. Some of these can be better reflected as an "or" to indicate that the paper doesn't have to be about both sub-topics. | Foodborne Illnesses or Food Safety |
| 34 | ['household water', 'drinking water', 'water sanitation', 'sanitation hygiene', 'water treatment', 'tap water', 'waterborne', 'water samples', 'water quality', 'sanitation'] | Drinking Water Quality | Water Quality | Water Safety and Hand Hygiene | water safety/contamination/pollution, sanitation and hygiene, water related outbreaks/risks/accidents, water fluoridation, handwashing, cholera | Water Safety and Hand Hygiene | Stage 2 reviewers requested this label reflect CDC language. Stage 2 reviewers requested clarifying any label where "and" occurs. Some of these can be better reflected as an "or" to indicate that the paper doesn't have to be about both sub-topics. | Water, Sanitation, and Hygiene |
| 35 | ['antifungal resistance', 'antifungal susceptibility', 'fungal infections', 'multidrug resistant', 'invasive fungal', 'fluconazole', 'antifungals', 'emerging multidrug', 'candidemia', 'fungal pathogen'] | Antifungal Resistance | Fungal Diseases | Fungal Infections--Epidemiolgic Trends and Treatment | fungal infections/illnesses, mycoses, fungi, molds, antifungals | Fungal Infections | | Fungal Infections |

| | | | | | | | | |
|---|---|---|---|---|---|---|---|---|
| 36 | ['child transmission', 'hiv infection', 'antenatal care', 'antiretroviral therapy', 'perinatal hiv', 'infant diagnosis', 'congenital syphilis', 'pregnant women', 'antenatal', 'syphilis screening'] | HIV Transmissi on | Mother-to-Child Transmissi on | Preventing Congenital Infections--HIV, Syphilis | congenital syphillis, congenital HIV, maternal/ne wborn screening for prevention/t hreats, mother child transmission | Congenital Infections | Stage 2 reviewers found it difficult to distinguish between this cluster and Viral Infections in Pregnancy. Most papers in this cluster are specifically about congenital syphilis and vertical transmission of HIV. | Vertical Transmission of HIV or Syphilis |
| 37 | ['sodium intake', 'health nutrition', 'nutrition', 'nutrition examination', 'ssb intake', 'dietary', 'diet', 'blood pressure', 'obesity', 'intakes'] | Nutrition in Diet | Nutritional Health | Diet, Health, Policy, & Disparities | nutrition, dietary intake/behav iors, food insecurity/ac cess, healthy eating, nutrition policies/mon itoring | Diet and Nutrition | Stage 2 reviewers requested listing nutrition first. Stage 2 reviewers requested clarifying any label where "and" occurs. Some of these can be better reflected as an "or" to indicate that the paper doesn't have to be about both sub-topics. | Nutrition and Diet |
| 38 | ['measles cases', 'measles surveillance', 'measles case', 'measles incidence', 'measles elimination', 'rubella elimination', 'measles outbreak', 'measles outbreaks', 'mumps outbreak', 'measles vaccination'] | Measles | Measles, Mumps, and Rubella | Vaccination & Vaccone Preventable Diseases of Children | measles, rubella, mumps, MMR vaccine | Measles, Mumps, and Rubella | | Measles, Mumps, and Rubella |

| | | | | | | | | |
|---|---|---|---|---|---|---|---|---|
| 39 | ['amebic meningoencephalitis', 'flaccid myelitis', 'meningoencephalitis', 'acute flaccid', 'myelitis', 'encephalitis', 'meningitis', 'cerebrospinal fluid', 'primary amebic', 'enterovirus'] | Rare and Fatal Infections in Children | Neuroinvasive Diseases | Infections Associated with Neurologic Symptoms, Immune Supression | viral etiology, outbreak etiology, unusual/emerging infectious diseases diagnosis/surveillance/investigations/detection, pathogenesis, central nervous system infections | Viral Infections with Neurological Symptoms | Stage 2 reviewers noted that some papers in this cluster presented parasitic infections, so requested the label be more generalized. | Infections with Neurological Symptoms |
| 40 | ['rabies prevention', 'rabies surveillance', 'rabies cases', 'canine rabies', 'rabies endemic', 'rabies elimination', 'rabies control', 'rabies exposure', 'rabies exposures', 'dog rabies'] | Rabies | Rabies | Global Rabies & Bat-Associated Pathogens | rabies, bat related disease/illnesses, animal related bites | Rabies | Stage 2 reviewers disagreed with the model on papers focused on bat-associated pathogens. | Rabies or Bat-Associated Pathogens |
| 41 | ['screening rates', 'screening uptake', 'cancer screening', 'breast cervical', 'cervical cancer', 'mammography use', 'national breast', 'screening test', 'cervical colorectal', 'breast cancer'] | Cancer Screening | Cancer Screenings | Cancer Prevention--Screening & Disparities | cancer screening/surveillance/detection/monitoring, cancer patient care, hysterectomy, mammography | Cancer Screening | Stage 2 reviewers noted some overlap with Cancer Epidemiology cluster. | Cancer Screening |

| | | | | | | | | |
|---|---|---|---|---|---|---|---|---|
| 42 | ['facepiece respirators', 'n95 respirators', 'respirators', 'respiratory protection', 'respirator', 'surgical masks', 'facepiece', 'healthcare workers', 'decontamination', 'filtering facepiece'] | Respiratory Protection | Masking as Respiratory Protection | Respiratory protection & Occupational Exposure Risks in Health Care | respirators, masks, PPE, respiratory protection/prevention, infection precautions, gowns/gloves, ventilation/air filtration, bloodborne pathogens, exposure control, occupational reproductive hazards | Respiratory Protection | Stage 2 reviewers noted presence of papers about gowns and other personal protective equipment (PPE) in this cluster. | Respiratory Protection or PPE |
| 43 | ['syringe services', 'immunodeficiency virus', 'hcv infection', 'inject drugs', 'hiv infection', 'drug use', 'persons inject', 'hepatitis virus', 'injection practices', 'injection drug'] | Bloodborne Pathogens | People Who Inject Drugs | Substance Abuse & Injection-Associated Infections | persons who inject drugs, illicit drug use, and risks for infectious disease in particular HIV/hepatitis | Substance Abuse and Associated Bloodborne Pathogens | Stage 2 reviewers requested clarifying any label where "and" occurs. Some of these can be better reflected as an "or" to indicate that the paper doesn't have to be about both sub-topics. | Substance Abuse and Associated Bloodborne Pathogens |
| 44 | ['ebola transmission', 'ebola cases', 'ebola outbreaks', 'disease ebola', 'ebola outbreak', 'cases ebola', 'virus disease', 'ebola virus', 'infection prevention', 'ebola epidemic'] | Ebola Virus | Ebola | Global Emergency Response -- EVD & Other Dangerous Pathogens | ebola, emergency operation centers, outbreak emergency response, public health detection and response mechanisms, COVID, anthrax, polio, nipah | Ebola and Global Emergency Response | Stage 2 reviewers requested clarifying any label where "and" occurs. Some of these can be better reflected as an "or" to indicate that the paper doesn't have to be about both sub-topics. | Ebola or Global Emergency Response |
| 45 | ['avian influenza', 'pathogenic avian', 'influenza hpai', 'influenza virus', 'influenza h7n9', 'influenza h5n1', 'influenza viruses', | Avian Influenza | Influenza Viruses | Avian Influenza-- Divergence, Interspecies Transmission | influenza | Pandemic Influenzas | Stage 2 reviewers requested pluralization change. | Pandemic Influenza |

| | | | | | | | | |
|---|---|---|---|---|---|---|---|---|
| | 'human infection', 'virus infection', 'human infections'] | | | | | | | |
| 46 | ['environmental health', 'environmental public', 'national environmental', 'environmental hazards', 'environmental', 'public health', 'biomonitoring', 'land reuse', 'exposures', 'health tracking'] | Environmental monitoring (combine 4 with 45) | Environmental Health | Environmental Health--Hazards, Tracking, Community | environmental health/exposures, toxic/hazardous substances, population exposures/biomonitoring, geospatial health | Environmental Health | Commonly mistaken for Chemical Exposures. Some representative papers in this cluster are about measurements in the actual environment, community initiatives, etc.; while representative papers in Chemical Exposures focused on measurements in people (urinary, blood serum). | Environmental Health |

Note: This table records additional context for topic clusters identified by the BERTopic modeling pipeline and records the review process that led to the assignment of final labels for each topic cluster. The “Topic Rank” column records the clusters in order from largest to smallest (i.e. the largest cluster has the Topic Rank of 1, while the smallest cluster has the Topic Rank of 46). The “Topic Model Keywords” column records the keywords the topic modeling pipeline returned that describe the papers in the topic cluster. The initial cluster labels assigned by “Reviewer A” and “Reviewer B” were based on review of the “Topic Model Keywords”. The initial cluster labels assigned by “Reviewer C” and “Reviewer D” were based on review of the titles and abstracts of papers in each cluster. The “Stage 1 Label” column reports the consensus of all four reviewers after discussion. The “Stage 2 Notes” column reports on additional discussion and revisions by Stage 2 reviewers, who were tasked with assigning sample papers to clusters based on the “Stage 1 Label” for the cluster. These notes give additional context and provide a justification for any revisions made to topic cluster labels. The “Final Label” reports the assigned label for each topic cluster after both stages of review by subject matter experts, incorporating any suggested revisions from Stage 2 reviewers.

**Supplementary Table 2.** *Number and percent of publications, median number of academic citations, % with any policy citations, median Altmetric Attention Score, academic metric rank, policy metric rank, and attention metric rank, for 46 public health topic themes identified by a large language model – CDC-authored publications, 2014-2023.*

| Public health topic theme | # | % | Median number of academic citations (2014-2020) | % with policy citations (2014-2020) | Median Altmetric Attention Score (2014-2023) | Academic metric rank | Policy metric rank | Attention metric rank |
|---|---|---|---|---|---|---|---|---|
| Respiratory Illnesses or Vaccination | 3828 | 11.2 | 22 | 63.3 | 10 | 9 | 9 | 3 |
| Sexually Transmitted Infections | 2931 | 8.6 | 14 | 40.7 | 3 | 36 | 36 | 33 |
| Occupational Safety and Health | 903 | 2.6 | 11 | 82.3 | 4 | 42 | 2 | 22 |
| Mining Safety and Health | 874 | 2.6 | 14 | 92.5 | 3 | 36 | 1 | 33 |
| Chemical Exposure Biomonitoring | 868 | 2.5 | 29 | 56.0 | 3 | 3 | 14 | 33 |
| Malaria or Parasitic Infections | 859 | 2.5 | 22 | 44.6 | 4 | 9 | 34 | 22 |
| Tuberculosis | 738 | 2.2 | 12 | 68.4 | 4 | 41 | 5 | 22 |
| Diabetes or Cardiovascular Health | 730 | 2.1 | 25 | 47.9 | 5 | 7 | 32 | 17 |
| Emerging or Zoonotic Infectious Diseases | 722 | 2.1 | 21 | 40.2 | 6 | 13 | 37 | 11 |
| Maternal and Child Health | 669 | 2.0 | 20 | 53.1 | 4 | 18 | 17 | 22 |
| Antimicrobial Resistance | 603 | 1.8 | 23 | 48.5 | 7 | 8 | 31 | 6 |
| Substance Abuse or Opioids | 587 | 1.7 | 29.5 | 64.2 | 14 | 2 | 8 | 1 |
| Violence | 577 | 1.7 | 27 | 62.1 | 5 | 4 | 10 | 17 |

| | | | | | | | | |
|---|---|---|---|---|---|---|---|---|
| Vaccine Development or Immune Response | 569 | 1.7 | 21 | 24.7 | 3 | 13 | 46 | 33 |
| Gastrointestinal Viruses | 560 | 1.6 | 20 | 32.5 | 2 | 18 | 42 | 45 |
| Tobacco or Smoking | 559 | 1.6 | 19.5 | 64.6 | 7 | 22 | 7 | 6 |
| Public Health Practice and Capacity | 538 | 1.6 | 7.5 | 41.8 | 3 | 44 | 35 | 33 |
| Nutritional Biomarkers | 525 | 1.5 | 21 | 51.1 | 3 | 13 | 25 | 33 |
| Injury | 522 | 1.5 | 17 | 52.9 | 6 | 27 | 19 | 11 |
| Viral Hepatitis | 494 | 1.4 | 17 | 51.4 | 3 | 27 | 24 | 33 |
| Non-Mosquito Vectorborne Infections | 484 | 1.4 | 20 | 32.5 | 4 | 18 | 42 | 22 |
| Mosquito and Vector Control | 472 | 1.4 | 20 | 39.0 | 3 | 18 | 39 | 33 |
| Global Health Security and Preparedness | 433 | 1.3 | 10 | 53.1 | 3 | 43 | 17 | 33 |
| Occupational Exposures | 400 | 1.2 | 18.5 | 79.7 | 6 | 23 | 4 | 11 |
| Cancer Epidemiology | 379 | 1.1 | 21 | 38.7 | 6 | 13 | 40 | 11 |
| Physical Activity | 365 | 1.1 | 18 | 39.9 | 4 | 24 | 38 | 22 |
| Polio | 353 | 1.0 | 17 | 51.9 | 5 | 27 | 21 | 17 |
| Intellectual and Developmental Health | 351 | 1.0 | 26 | 66.7 | 7 | 5 | 6 | 6 |
| Human Papillomavirus | 345 | 1.0 | 18 | 50.8 | 4 | 24 | 26 | 22 |
| Pneumococcal, Meningococcal, or Streptococcal Infections | 345 | 1.0 | 22 | 50.4 | 4 | 9 | 27 | 22 |

| | | | | | | | | |
|---|---|---|---|---|---|---|---|---|
| Microbial Genomics of Foodborne Pathogens | 327 | 1.0 | 16 | 28.9 | 3 | 31 | 45 | 33 |
| Viral Infections and Pregnancy | 307 | 0.9 | 25.5 | 60.5 | 11 | 6 | 12 | 2 |
| Foodborne Illnesses or Food Safety | 304 | 0.9 | 22 | 61.2 | 8 | 9 | 11 | 5 |
| Water, Sanitation, and Hygiene | 293 | 0.9 | 13 | 46.6 | 3 | 38 | 33 | 33 |
| Fungal Infections | 293 | 0.9 | 36.5 | 36.0 | 10 | 1 | 41 | 3 |
| Vertical Transmission of HIV or Syphilis | 282 | 0.8 | 13 | 53.7 | 3 | 38 | 16 | 33 |
| Nutrition and Diet | 279 | 0.8 | 16 | 50.2 | 6 | 31 | 28 | 11 |
| Measles, Mumps, and Rubella | 275 | 0.8 | 15 | 51.6 | 7 | 34 | 22 | 6 |
| Infections with Neurological Symptoms | 240 | 0.7 | 17 | 31.9 | 4 | 27 | 44 | 22 |
| Rabies or Bat-Associated Pathogens | 230 | 0.7 | 18 | 54.3 | 6 | 24 | 15 | 11 |
| Cancer Screening | 214 | 0.6 | 15 | 49.1 | 5 | 34 | 30 | 17 |
| Respiratory Protection or PPE | 204 | 0.6 | 13 | 79.9 | 4 | 38 | 3 | 22 |
| Substance Abuse and Associated Bloodborne Pathogens | 184 | 0.5 | 15.5 | 57.9 | 4 | 33 | 13 | 22 |
| Ebola / Global Emergency Response | 181 | 0.5 | 6.5 | 50.0 | 5 | 45 | 29 | 17 |
| Pandemic Influenza | 177 | 0.5 | 20.5 | 52.8 | 7 | 17 | 20 | 6 |
| Environmental Health | 175 | 0.5 | 6.5 | 51.6 | 2 | 45 | 22 | 45 |
| ***Total – Clustered into Themes*** | 26548 | 77.8 | 18 | 53.3 | 4 | -- | -- | -- |

| | | | | | | | | |
|---|---|---|---|---|---|---|---|---|
| ***Total - Unclustered*** | 7556 | 22.2 | 17 | 49.4 | 4 | -- | -- | -- |
| ***Total*** | 34104 | 100 | 18 | 52.4 | 4 | -- | -- | -- |

Note: Topics are ordered from the largest to smallest cluster. For measures of academic and policy impact, only publications that are at least 3 years old are included, since these have had time to accumulate citations (See Supplemental Figure 1). The “Median Academic Citations (2014-2020)” column reports the median number of academic citations for papers in the cluster as a measure of academic impact. The “% with Policy Citations (2014-2020)” column reports the percent of publications in the cluster with at least one academic citation as a measure of policy impact. The “Median AAS (2014-2023)” column reports the median Altmetric Attention Score (AAS) for publications in the cluster as a measure of attention impact. The “Academic Rank”, “Policy Rank”, and “Attention Rank” columns report how clusters rank for academic, policy, and attention impact, respectively (i.e. the highest impact cluster has a rank of 1, and the lowest impact cluster has a rank of 46; clusters can tie in rank). For each topic, their highest impact rank across the three clusters is highlighted in gray.
